# mRNA-based COVID-19 vaccine boosters induce neutralizing immunity against SARS-CoV-2 Omicron variant

**DOI:** 10.1101/2021.12.14.21267755

**Authors:** Wilfredo F. Garcia-Beltran, Kerri J. St. Denis, Angelique Hoelzemer, Evan C. Lam, Adam D. Nitido, Maegan L. Sheehan, Cristhian Berrios, Onosereme Ofoman, Christina C. Chang, Blake M. Hauser, Jared Feldman, David J. Gregory, Mark C. Poznansky, Aaron G. Schmidt, A. John Iafrate, Vivek Naranbhai, Alejandro B. Balazs

## Abstract

Recent surveillance has revealed the emergence of the SARS-CoV-2 Omicron variant (BA.1/B.1.1.529) harboring up to 36 mutations in spike protein, the target of vaccine-induced neutralizing antibodies. Given its potential to escape vaccine-induced humoral immunity, we measured neutralization potency of sera from 88 mRNA-1273, 111 BNT162b, and 40 Ad26.COV2.S vaccine recipients against wild type, Delta, and Omicron SARS-CoV-2 pseudoviruses. We included individuals that were vaccinated recently (<3 months), distantly (6-12 months), or recently boosted, and accounted for prior SARS-CoV-2 infection. Remarkably, neutralization of Omicron was undetectable in most vaccinated individuals. However, individuals boosted with mRNA vaccines exhibited potent neutralization of Omicron only 4-6-fold lower than wild type, suggesting that boosters enhance the cross-reactivity of neutralizing antibody responses. In addition, we find Omicron pseudovirus is more infectious than any other variant tested. Overall, this study highlights the importance of boosters to broaden neutralizing antibody responses against highly divergent SARS-CoV-2 variants.

## INTRODUCTION

The SARS-CoV-2 Omicron variant (BA.1/B.1.1.529) was first detected in Botswana and reported to the World Health Organization (WHO) in November 2021 as a novel variant with an unprecedented number of previously described and novel mutations with immunevasive potential. A subsequent and rapid increase in Omicron cases in South Africa resulted in its designation as a novel variant of concern (VOC) by the World Health Organization (WHO) (www.who.int). This variant harbors up to 59 mutations throughout its genome, with as many as 36 of these occurring within the spike protein, the mediator of host cell entry and the main target of neutralizing antibodies. Studies of previous SARS-CoV-2 variants have demonstrated that mutations within the receptor binding domain (RBD) mediate escape from vaccine-induced neutralizing antibodies (Cele et al., 2021a; Garcia-Beltran et al., 2021a; Zhou et al., 2021) and in some cases increase infectivity through enhanced affinity for ACE2 (Tian et al., 2021). Omicron RBD contains 15 mutations, some of which overlap with previously studied variants. For example, Beta (B.1.351) and Gamma (P.1) harbor mutations in residues K417, E484, and N501 that potently diminish vaccine-induced neutralization (Garcia-Beltran et al., 2021a), possibly the result of neutralizing antibody responses being focused towards a limited set of RBD epitopes, as has been previously described (Barnes et al., 2020; Greaney et al., 2021a).

In the United States, three vaccines have been approved by the FDA or are under emergency use authorization (EUA), all of which use the original wild type SARS-CoV-2 spike protein first identified in Wuhan, China as the sole immunogen. These are formulated as spike-encoding mRNA in lipid nanoparticles (BNT162b2 manufactured by Pfizer-BioNTech and mRNA-1273 manufactured by Moderna) or as an adenovirus vectored vaccine (Ad26.COV2.S manufactured by Janssen/Johnson & Johnson) (Baden et al., 2021; Polack et al., 2020; Sadoff et al., 2021). These SARS-CoV-2 vaccines have been remarkably successful in inducing neutralizing humoral and cellular immunity and more importantly, reducing COVID-19 infections, hospitalizations, and deaths in clinical trials (Baden et al., 2021; Polack et al., 2020; Sadoff et al., 2021) and during rapid worldwide deployment (Tregoning et al., 2021). However, it has now been shown that neutralizing antibody responses and vaccine effectiveness vary by vaccine agent, decrease with increased time post vaccination, and are negatively impacted by emerging variants (Bajema et al., 2021; Bar-On et al., 2021; Cromer et al., 2021; Khoury et al., 2021; Lopez Bernal et al., 2021; Naranbhai et al., 2021a; Tregoning et al., 2021). In an effort to combat waning antibody responses and the emergence of new variants, boosters have been approved for individuals vaccinated >6 months ago and have been shown to be very effective at inducing high neutralizing antibody titers (Bar-On et al., 2021). However, while neutralization of wild type SARS-CoV-2 has been shown to predict the effectiveness of vaccines against variants (Earle et al., 2021; Khoury et al., 2021), it is unclear whether this correlation will be maintained among boosters and for highly mutated variants like Omicron (Bajema et al., 2021; Lopez Bernal et al., 2021; Tregoning et al., 2021).

We previously developed and validated a high-throughput pseudovirus neutralization assay to understand differences in immunity by vaccine and host characteristics against SARS-CoV-2 variants and other coronaviruses (Garcia-Beltran et al., 2021a, 2021b; Naranbhai et al., 2021a). Here, we used this assay to test sera from 239 individuals who had been fully-vaccinated with one of three vaccines approved in the United States—mRNA-1273, BNT162b2, or Ad26.COV2.S—against wild type, Delta, and Omicron SARS-CoV-2 pseudoviruses. This included 70 individuals who had received an mRNA vaccine booster >6 months after primary vaccination series as either a cross-over or solely mRNA-based vaccination regimen. Remarkably, we found that all three primary vaccine series resulted in low-to-absent neutralization of SARS-CoV-2 Omicron. However, boosted mRNA vaccine recipients exhibited potent neutralization against Omicron, despite exhibiting wild type neutralization titers similar to those in recently vaccinated (non-boosted) individuals. In addition, *in-vitro* infectivity experiments demonstrated that Omicron pseudovirus continues to rely upon the human ACE2 receptor for host cell entry and is nearly 4-fold more infectious than wild-type pseudovirus and 2-fold more infectious than Delta pseudovirus. Together, our results highlight that SARS-CoV-2 Omicron variant evades vaccine-induced neutralizing immunity under current vaccine regimens and is more infectious than previous variants. Notwithstanding, our finding of potent cross-neutralizing immunity against Omicron in boosted individuals suggests that existing mRNA vaccines may overcome evasion of humoral immunity by future variants of concern.

## RESULTS

### Emergence of SARS-CoV-2 Omicron as a novel and highly mutated variant of concern

Over the course of more than 270 million confirmed SARS-CoV-2 infections worldwide, the virus has undergone remarkable diversification, producing >1,500 uniquely identified Pango lineages (Rambaut et al., 2020) (coronavirus.jhu.edu). Some of these have demonstrated evidence of increased transmissibility, virulence, and/or immune evasion, prompting the WHO to classify five lineages as current VOCs (www.who.int). The Omicron variant, also known as PANGO lineage BA.1 or B.1.1.529, was first reported in November 2021 and received its VOC designation within days on account of its unique mutational profile and dramatic rise in cases observed in Gauteng, South Africa. While the Delta variant is now the dominant SARS-CoV-2 variant worldwide after overtaking the Alpha variant in July 2021, the rise of Omicron infections in regions where Delta is circulating suggests that Omicron may overtake Delta to become the next dominant strain. Despite the substantial recent expansion of the Delta lineage, phylogenetic analysis suggests that the Omicron variant was derived from the Alpha lineage and only recently detected by genomic surveillance (**Figure 1A**). In comparison to the 9 mutations or deletions found in Delta, the Omicron lineage we tested harbors 34 mutations (including three deletions and one insertion) in the spike protein including 15 within the RBD region (**Figure 1B**). These mutations are structurally focused at the top of the spike, in regions accessible to antibodies, raising the likelihood of immune evasion (**Figure 1C**).

**Figure 1.**
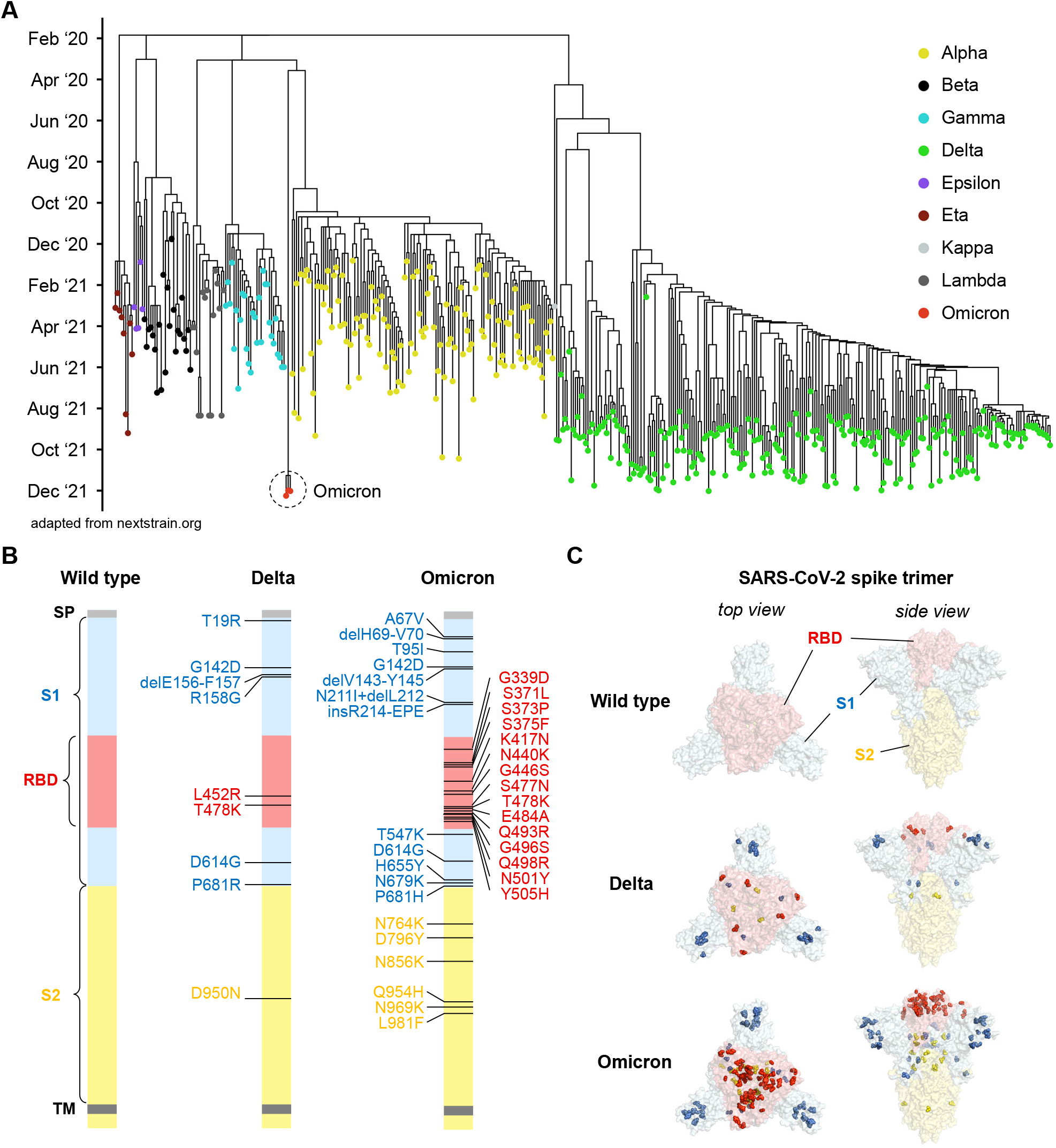
Emergence of SARS-CoV-2 Omicron among global variants of concern. **(A)** Phylogenetic tree of SARS-CoV-2 variants with sampling dates shows emergence of Omicron variant by December 2021 (adapted from nextstrain.org). **(B)** Schematic of SARS-CoV-2 spike protein structure and mutations of variants used in this study are illustrated. Omicron variant mutations used in this study were based on the most prevalent mutations (>85% frequency) found in GISAID and reflect the dominant Omicron variant. The regions within the spike protein are abbreviated as follows: SP, signal peptide; RBD, receptor binding domain; TM, transmembrane domain. **(C)** Crystal structure of pre-fusion stabilized SARS-Cov-2 spike trimer (PDB ID 7JJI) highlighting the mutational landscape of SARS-CoV-2 Delta and Omicron variants relative to SARS-CoV-2 wild type. Top views (*left panels*) and side views (*right panels*) of spike protein are shown with mutations in RBD (in red), S1 (in blue), and S2 (in yellow) highlighted with residue atoms as colored spheres.

### Neutralizing antibody responses to SARS-CoV-2 variants demonstrate substantial escape by Omicron

We accrued a diverse cohort of 239 COVID-19 vaccinees that were healthcare workers and/or community dwellers from Boston or Chelsea, Massachusetts (**Supplementary Table S1**). The entire cohort had a median age of 38 years (range: 18 - 78 years) and was 63% female. Vaccinees had received a full series of mRNA-1273, BNT162b, or Ad26.COV2.S and were subdivided into the following subgroups: infection-naive individuals that received their primary vaccination series within the last 3 months (‘recent vax’); infection-naive individuals that received their primary vaccination series 6 – 12 months before (‘distant vax’); individuals that received their primary vaccination series 6 – 12 months before and had a history of self-reported positive PCR and/or serologic evidence of SARS-CoV-2 infection as measured by anti-nucleocapsid antibodies (‘distant vax + infection’); and infection-naive individuals that were boosted within the last 3 months (‘booster vax’) (**Figure 2A**).

**Figure 2:**
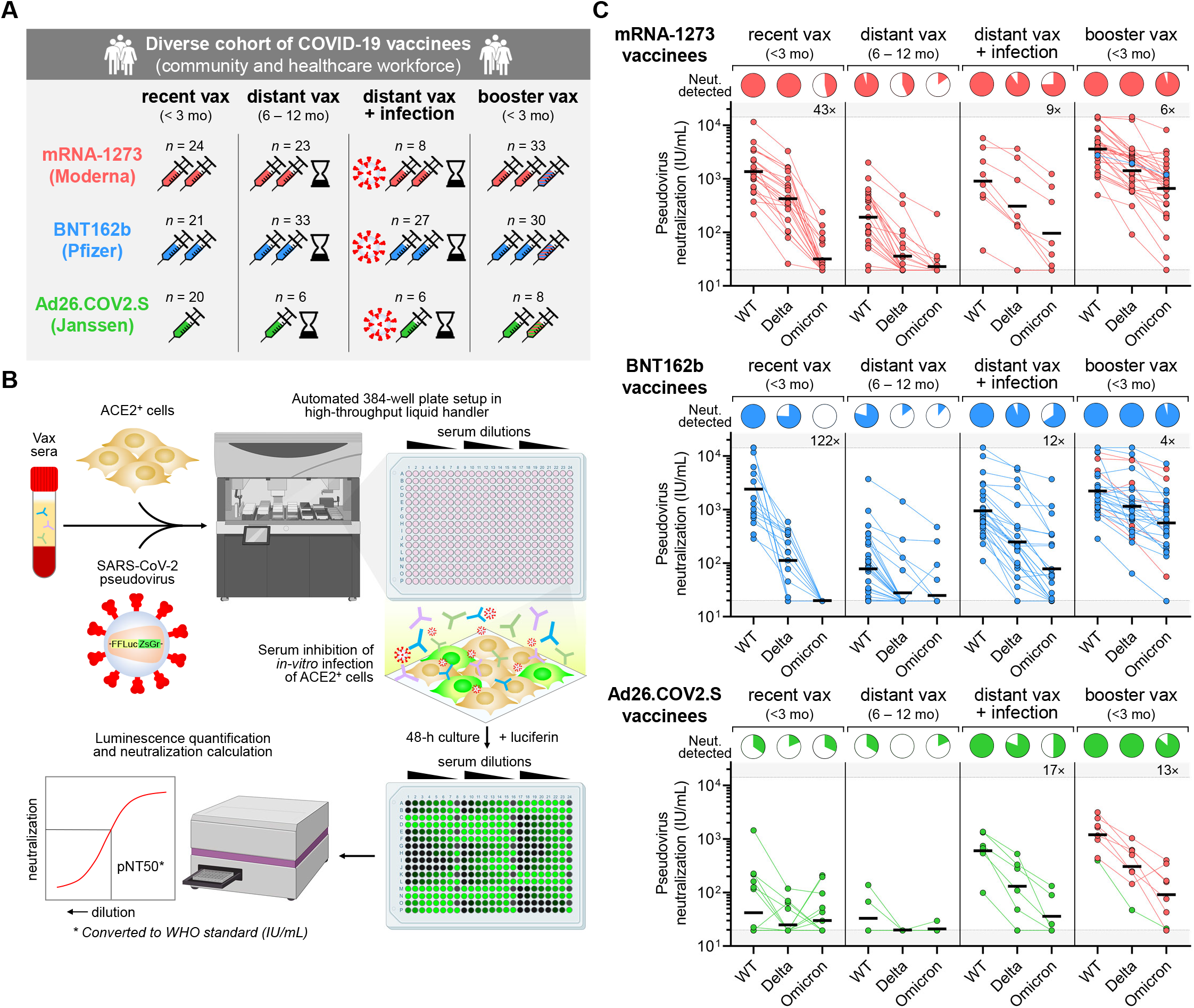
mRNA vaccine booster induces potent neutralizing responses against SARS-CoV-2 Omicron variant that are low-to-absent in non-boosted vaccinees. **(A)** Schematic representation of vaccinee cohorts of healthy adult community dwellers and health care workers. Participants who completed their primary series of vaccination with two-dose mRNA-1273 (Moderna), two-dose BNT162b (Pfizer), or 1-dose Ad26.COV2.S (Janssen) were included in this study. Vaccinees were stratified into four subgroups as follows: infection-naive, non-boosted individuals that received primary vaccination series within last 3 months (‘recent vax’); individuals that received primary vaccination series 6 – 12 months before and were either without (‘distant vax’) or with a history or serologic evidence of SARS-CoV-2 infection (‘distant vax + infection’); and infection-naive individuals that were boosted within the last 3 months (‘booster vax’). History of SARS-CoV-2 infection was determined by either self-reported history of positive PCR test and/or positive anti-SARS-CoV-2 nucleocapsid antibody test. **(B)** Schematic of experimental workflow of high-throughput SARS-CoV-2 pseudovirus neutralization assay used to determine neutralization titer of vaccinee sera against variants. **(C)** Neutralization titers (in WHO IU/mL) of wild type (WT), Delta, and Omicron pseudoviruses were determined for people who received primary vaccination series with mRNA-1273 (*top panel*; in red), BNT162b2 (*middle panel*; in blue), or Ad26.COV2.S (*bottom panel*; in green) and classified into the aforementioned subgroups (in **A**). Dark horizontal lines for each group denote geometric mean titer. Pie charts show the proportion of vaccinees within each group that had detectable neutralization against the indicated SARS-CoV-2 pseudovirus. Fold-decrease in geometric mean neutralization titer of Omicron relative to wild type within a subgroup is shown as a number with ‘×’ symbol within the gray region; this was done only for vaccinee subgroups where neutralization against wild type pseudovirus was detected in 100% of individuals. All fold-decreases shown have unadjusted *p* < 0.05 with paired *t* test. Within ‘booster vax’ subgroups (*far right*), boosters were homologous (same vaccine) except for 1 of 33 mRNA-1273 vaccinees that crossed-over to BNT162b (*top panel*; in blue), 6 of 30 BNT162b vaccinees that crossed-over to mRNA-1273 (*middle panel*; in red), and 7 of 8 Ad26.COV2.S vaccinees crossed-over to mRNA-1273 (*bottom panel*; in red).

Sera from these cohorts were subjected to a high-throughput luminescence-based neutralization assay that we and others have previously validated (Crawford et al., 2020; Garcia-Beltran et al., 2021b; Ju et al., 2020; Moore et al., 2004; Pinto et al., 2020; Wang et al., 2020; Yang et al., 2020) to assess SARS-CoV-2 variant neutralization (**Figure 2B**). In brief, pseudovirus encoding luciferase and bearing SARS-CoV-2 variant spike proteins were exposed to dilutions of vaccinee sera prior to being added to ACE2-expressing target cells. After 48 h of co-culture, luciferase activity of the dilution series was measured to quantify infection rates and calculate the titer that achieved 50% pseudovirus neutralization (pNT50). This pNT50 value was subsequently converted to WHO IU/mL after correcting with a WHO reference standard that was run in parallel (Knezevic et al., 2021).

In line with previous studies (Naranbhai et al., 2021a, 2021b), individuals that were recently vaccinated with mRNA-1273 or BNT162b achieved substantially higher wild type neutralization titers than Ad26.COV2.S vaccinees, with geometric mean neutralization titers (GMNT) of 1,362 IU/mL for mRNA-1273, 2,402 IU/mL for BNT162b, and 42 IU/mL for Ad26.COV2.S (**Figure 2C**). Individuals vaccinated >6 months prior exhibited substantially lower but mostly detectable wild type neutralization (GMNT 192 IU/mL for mRNA-1273, 78 IU/mL for BNT162b, and 33 IU/mL for Ad26.COV2.S) (**Figure 2C**). Prior history of infection was associated with high levels of wild type neutralization titers even in distantly vaccinated individuals, particularly in Ad26.COV2.S vaccinees (GMNT 904 IU/mL for mRNA-1273; 947 IU/mL for BNT162b; and 603 IU/mL for Ad26.COV2.S) (**Figure 2C**). However, recently boosted individuals exhibited among the highest neutralization titers against wild type SARS-CoV-2 pseudovirus (GMNT 3,862 IU/mL for mRNA-1273; 2,219 IU/mL for BNT162b; and 1,201 IU/mL for Ad26.COV2.S) (**Figure 2C**).

Neutralization of Delta variant pseudovirus was decreased relative to wild type for all subgroups (**Figure 2C**), as has been previously reported (Mlcochova et al., 2021; Planas et al., 2021). While Delta neutralization became undetectable in most individuals vaccinated >6 months before blood draw, Delta neutralization was detectable and only modestly decreased in recently vaccinated, previously infected, and recently boosted vaccinees (**Figure 2C**). In contrast, Omicron neutralization was dramatically decreased among all subgroups, including recently vaccinated mRNA-1273 and BNT162b recipients, which demonstrated a complete loss of neutralization in >50% of individuals and GMNT decrease of 43-fold for mRNA-1273 and 122-fold for BNT162b (**Figure 2C**). Previously infected vaccinees also had a substantial decrease in Omicron neutralization titer (GMNT decrease of 9-fold for mRNA-1273, 12-fold for BNT162b, and 17-fold for Ad26.COV2.S), but most retained detectable neutralization (**Figure 2C**). Remarkably, however, recently boosted vaccinees exhibited potent neutralization of Omicron variant pseudovirus that was only moderately decreased relative to wild type neutralization (GMNT decrease of 6-fold for mRNA-1273, 4-fold for BNT162b, and 13-fold for Ad26.COV2.S) (**Figure 2C**). Of note, among boosted vaccinees, all boosters were homologous (same vaccine) except for 1 of 33 mRNA-1273 vaccinees that crossed-over to BNT162b, 6 of 30 BNT162b vaccinees that crossed-over to mRNA-1273, and 7 of 8 Ad26.COV2.S vaccinees that crossed-over to mRNA-1273. Taken together, our results indicate that two-dose mRNA-based vaccines are effective at inducing neutralizing immunity to SARS-CoV-2 wild type and Delta variants but suboptimal for inducing neutralizing responses to the Omicron variant.

### mRNA vaccine boosters increase breadth and cross-reactivity of neutralizing antibody response

Given the drastic increase in cross-neutralization of SARS-CoV-2 Omicron pseudovirus in boosted versus non-boosted vaccinees, we directly compared sera from individuals that recently received their primary series to those that were boosted with an mRNA vaccine within the last 3 months. Wild type pseudovirus neutralization was comparable between individuals who received three versus two doses of either mRNA vaccine (GMNT increase of 3-fold for mRNA-1273 and 1-fold for BNT162b). The difference in neutralization of Delta pseudoviruses between three-dose and two-dose vaccinees was similar for mRNA-1273 (GMNT increase of 3-fold) but increased in BNT162b vaccinees (GMNT increase of 9-fold). Cross-neutralization of Omicron variant was substantially higher in individuals who received three doses of either mRNA vaccine (GMNT increase of 19-fold for mRNA-1273 and 27-fold for BNT162b) (**Figure 3A**). Interestingly, Ad26.COV2.S vaccinees boosted with mRNA-1273 showed substantially higher wild type, Delta, and Omicron pseudovirus neutralization relative to those who received Ad26.COV2.S alone (**Figure 3A**).

**Figure 3:**
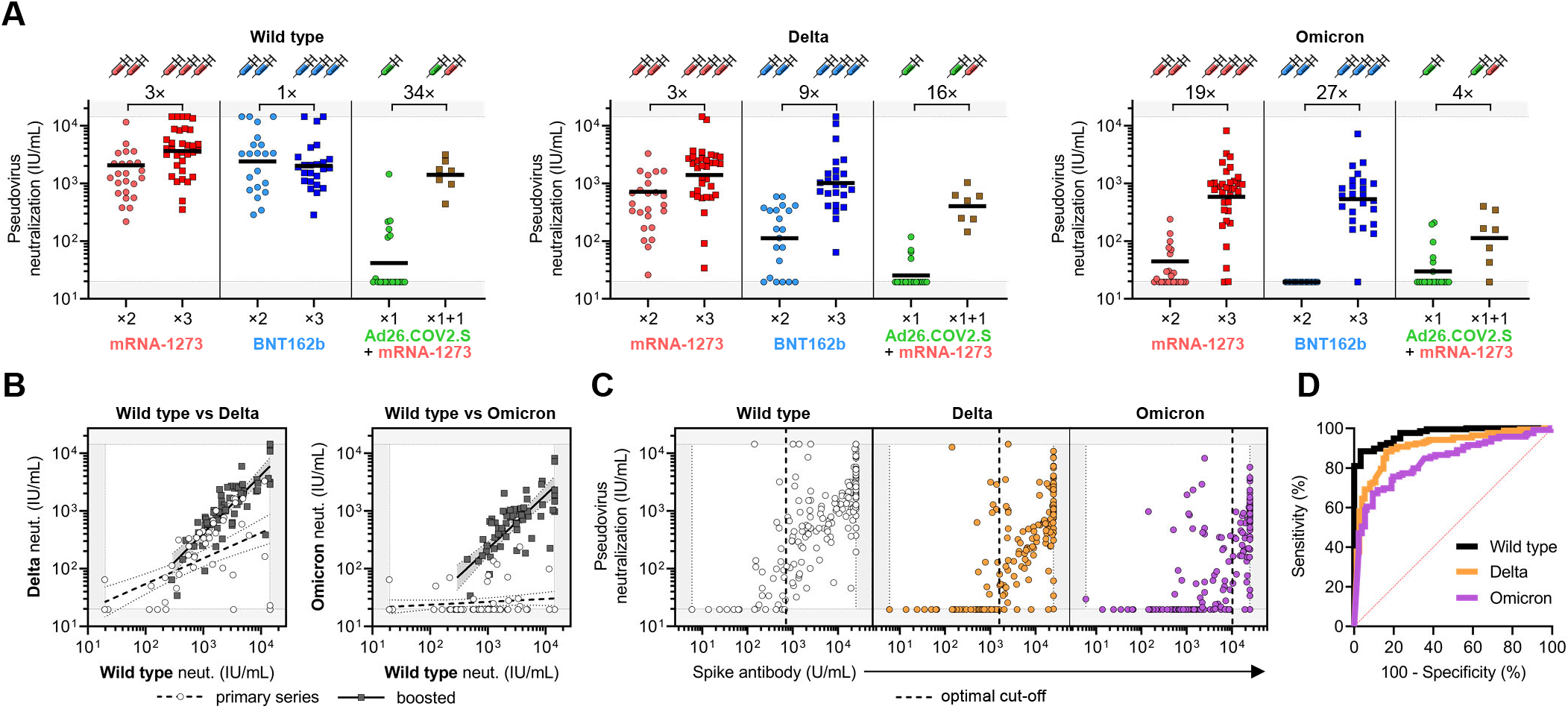
Cross-reactivity of neutralizing antibody response is increased by mRNA vaccine booster relative to primary vaccination series and can be predicted by anti-spike antibody levels. **(A)** Neutralization titers (in WHO IU/mL) of wild type (WT; *left pane*l), Delta (*middle panel*), and Omicron (*right panel*) SARS-CoV-2 pseudoviruses were analyzed for infection-naive participants that were recently vaccinated with primary series or booster (<3 months). Recently vaccinated individuals received mRNA-1273 (×2), BNT162b (×2), or Ad26.COV2.S (×1) and boosted individuals received a homologous booster of mRNA-1273 (×3) or BNT162b (×3) or a cross-over booster of mRNA-1273 for Ad26.COV2.S vaccinees (×1+1). Fold-increase in geometric mean neutralization titer of boosted versus non-boosted individuals is shown as a number with ‘×’ symbol. This analysis is based on experimental data depicted in **Figure 2C** but excludes participants with prior SARS-CoV-2 infection, distant vaccination (>6 months), and/or cross-over between mRNA-1273 and BNT162b to understand differences in neutralizing responses soon after primary vaccination series versus boosting. The single Ad26.COV2.S vaccinee that received a homologous boost with Ad26.COV2.S was also excluded from this analysis. Dark horizontal lines for each group denote geometric mean titer. **(B)** Aggregate data from study participants in **A** that recently received primary vaccination series (‘primary series’; white circles) or were recently boosted (‘boosted’; dark gray squares) was used for linear regression analysis of wild type versus Delta (*left panel*) or wild type versus Omicron (*right panel*) pseudovirus neutralization. Wild type neutralization titers correlated with Delta neutralization in ‘primary series’ individuals (*R*^2^ = 0.35; slope = 0.44; *p* < 0.0001), and even more strongly in ‘boosted’ individuals (*R*^2^ = 0.68; slope = 1.00; *p* < 0.0001). Wild type neutralization titers showed no significant relationship with Omicron neutralization in ‘primary series’ individuals (*R*^2^ = 0.03; slope = 0.05; *p* = 0.16); however, ‘boosted’ individuals showed a significant correlation with Omicron neutralization titers (*R*^2^ = 0.56; slope = 0.94; *p* < 0.0001). **(C)** Anti-SARS-CoV-2 spike antibodies levels (measured by EUA-approved clinical diagnostic test) of all vaccinees were plotted against neutralization of wild type (*left panel*; white circles), Delta (*middle panel*; orange circles), and Omicron (*right panel*; purple circles) SARS-CoV-2 pseudoviruses. Optimal spike antibody cut-offs for predicting positive neutralization were determined by ROC analyses in **D** and are indicated with a vertical dashed line. **(D)** Receiver operating characteristic (ROC) analyses assessing the ability of spike antibody levels to predict neutralization of wild type (black line), Delta (orange line), and Omicron (purple line) pseudoviruses. Positive neutralization was defined as >33 IU/mL (previously defined with a cohort of 1,200 pre-pandemic samples (Garcia-Beltran et al., 2021b) and converted to WHO IU/mL). Area under curve (AUC) for wild type was 0.97, Delta was 0.91, and Omicron was 0.84, with *p* < 0.0001 for all three. Optimal cut-offs that maximized sensitivity (Se) and specificity (Sp) were determined using the ‘Se + Sp’ method, and were as follows: for wild type, optimal cut-off of 711 U/mL achieved 88.4% Se and 96.7% Sp; for Delta, optimal cut-off of 1,591 U/mL achieved 88.4% Se and 83.8% Sp; and for Omicron variant, optimal cut-off of 10,300 achieved 67.2% Se and 90.6% Sp. These optimal cut-off values are plotted as a vertical dashed line in **C**.

To better characterize the neutralization patterns observed between individuals who were fully vaccinated with each of the three approved vaccines and those who were boosted, we directly compared the wild type neutralization activity of these two groups of samples against Delta and Omicron pseudovirus (**Figure 3B**). Interestingly, we found that wild type neutralization titers from individuals who received their primary series correlated weakly to Delta variant cross-neutralization (*R*^2^ = 0.35; slope = 0.44; *p* < 0.0001), and did not correlate with Omicron variant cross-neutralization (*R*^2^ = 0.03; slope = 0.05; *p* = 0.16). In contrast, wild type neutralization of boosted individuals correlated strongly with Delta (*R*^2^ = 0.68; slope = 1.00; *p* < 0.0001) and Omicron (*R*^2^ = 0.56; slope = 0.94; *p* < 0.0001) variant cross-neutralization. This indicates that in addition to inducing higher neutralization titers against wild type SARS-CoV-2, boosting increases the breadth of humoral immunity and cross-reactivity against highly mutated SARS-CoV-2 variants such as Omicron.

### Neutralization of SARS-CoV-2 variants can be predicted by anti-spike antibody levels

Given the widespread accessibility of anti-SARS-CoV-2 spike serological assays in the clinical setting, we correlated the neutralization signal obtained by our high-throughput assay to measurements made by the Roche Elecsys® anti-SARS-CoV-2 spike semi-quantitative immunoassay in the clinical laboratory. As expected, we observed higher neutralization of wild type, Delta, and Omicron pseudoviruses with higher levels of anti-spike antibodies among all vaccinees (**Figure 3C**). Receiver operating characteristic (ROC) analysis was performed to assess how well anti-spike antibody levels performed at predicting positive neutralization of wild type, Delta, and Omicron pseudoviruses, which was defined as neutralization titer >33 IU/mL (based on previously established cohort of 1,200 pre-pandemic samples (Garcia-Beltran et al., 2021b) and converted to WHO IU/mL). Area under curve (AUC) for wild type was 0.97, Delta was 0.91, and Omicron was 0.84, and the optimal cut-offs that maximized sensitivity (Se) and specificity (Sp) were determined using the ‘Se + Sp’ method, resulting in following: for wild type, optimal cut-off of 711 U/mL achieved 88.4% Se and 96.7% Sp; for Delta, optimal cut-off of 1,591 U/mL achieved 88.4% Se and 83.8% Sp; and for Omicron variant, optimal cut-off of 10,300 achieved 67.2% Se and 90.6% Sp (**Figure 3C** and **3D**). This highlights the potential use of this widely available clinical diagnostic test in predicting SARS-CoV-2 variant neutralization.

### *Omicron retains ACE2 usage and exhibits higher infectivity than other SARS-CoV-2 variants* in vitro

Having detected a substantial degree of evasion of vaccine-induced humoral responses by Omicron, we next investigated how mutations in Omicron might result in changes in infectivity compared to wild type SARS-CoV-2. To investigate whether the spike of circulating variants mediate host cell entry via ACE2, we examined the ability of pseudoviruses bearing wild type, Delta, and Omicron spike to infect 293T-ACE2 cells or parental 293T cells devoid of ACE2 receptor (**Figure 4A**). None of the pseudoviruses tested infected 293T cells in the absence of ACE2 protein, confirming that like other SARS-CoV-2 strains, Omicron remains dependent on ACE2 for host cell entry (**Figure 4B**). To comprehensively determine quantitative differences in the infectivity of SARS-CoV-2 variants of concern, we compared the efficiency of a panel of pseudovirus variants to infect 293T-ACE2 cells over a range of viral concentrations. Remarkably, Omicron pseudovirus exhibited greater infection of target cells regardless of concentration when compared to all other tested variants (**Figure 4C**). Comparison of the linear regressions of each pseudovirus to wild type over the entire range revealed that, whereas Gamma variant exhibited similar infectivity to wild type, Beta was less infectious, and Delta was nearly 2-fold more infectious. Strikingly, Omicron was 4-fold more infectious than wild type and 2-fold more infectious than Delta. Taken together, these data demonstrate distinct differences in infectivity according to spike sequence, with Omicron exhibiting more efficient ACE2-mediated infection than wild type or other variant strains (**Figure 4D**).

**Figure 4.**
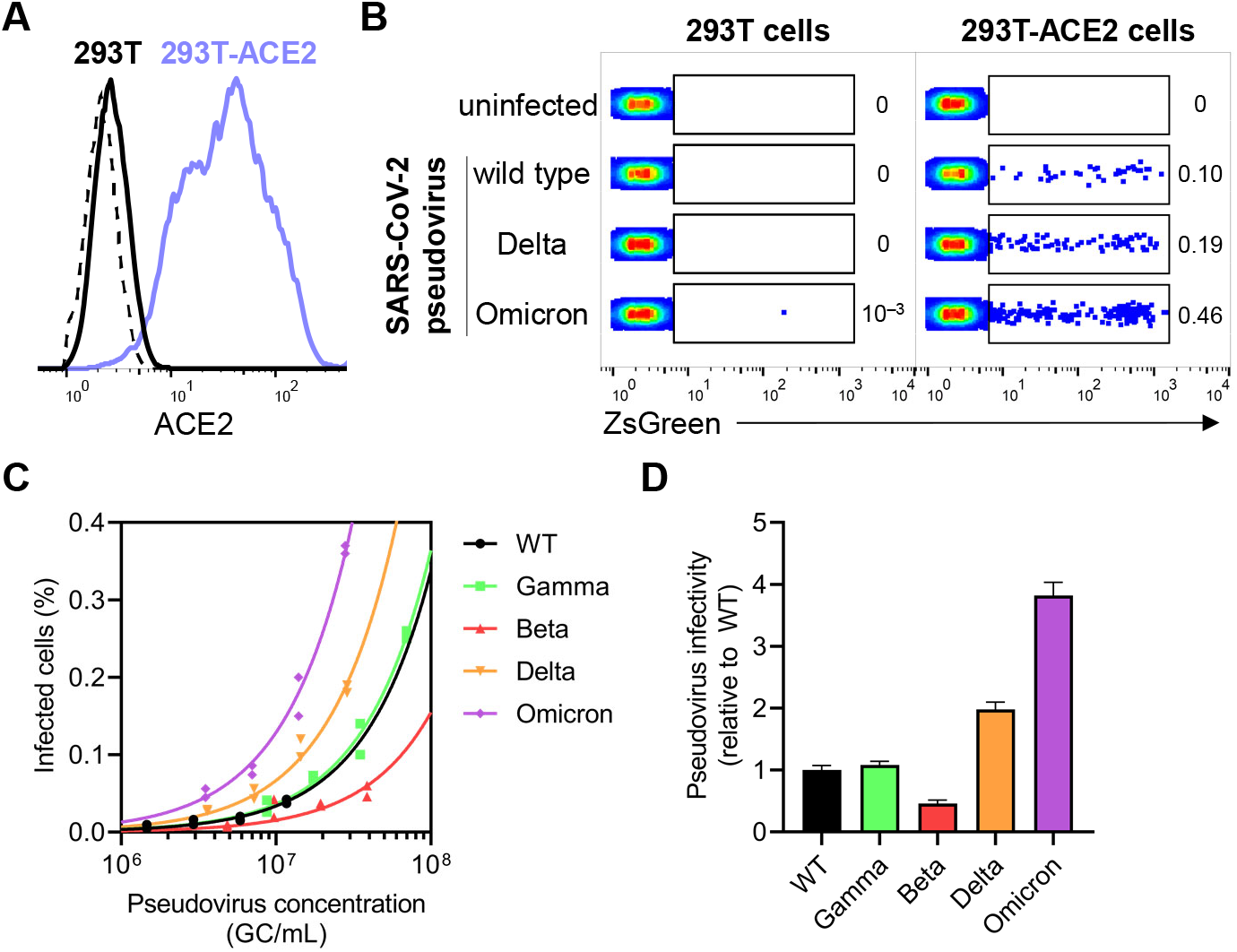
SARS-CoV-2 Omicron pseudovirus demonstrates a substantial increase in infectivity of ACE2^+^ cells relative to other SARS-CoV-2 variants *in vitro*. **(A)** Flow cytometry histogram depicting ACE2 surface staining on parental 293T cell line (in black), and 293T-ACE2 cell line stably expressing human ACE2 (in lilac). Dashed line indicates unstained control. **(B)** Representative dot plots show percentage of 293T (*left panel*) and 293T-ACE2 (*right panel*) cells that were infected with wild type, Delta, or Omicron SaRS-CoV-2 pseudoviruses (within ZsGreen^+^ gate) after 48 h of co-culture. Pseudoviruses were produced in parallel and under identical conditions to ensure comparable titers. **(C)** Titering of SARS-CoV-2 pseudoviruses of wild type, Gamma, Beta, Delta, and Omicron variants was performed on 293T-ACE2 cells and correlated to pseudovirus concentration in genome copies (GC)/mL as determined by qPCR. Two technical replicates (*n* = 2) were performed and a linear regression was performed to fit data given that a linear relationship of virus concentration versus infected cells can be assumed at infection rates <10%. **(D)** Pseudovirus infectivity relative to wild type was measured for each SARS-CoV-2 variant in **C** by calculating fold change in slope from **C** for each pseudovirus relative to wild type. Bars and error bars depict mean and standard error of the mean.

## DISCUSSION

Prior characterization of vaccine-induced humoral immunity against SARS-CoV-2 variants of concern revealed significant loss of activity against the Gamma and Beta variants, owing largely to three mutations in the RBD region of spike (Garcia-Beltran et al., 2021a). Given the 15 mutations present in the RBD of Omicron, which overlap the three sites mutated in Gamma and Beta (K417, E484, N501), as well as prior work done to understand the potential for antibody escape (Schmidt et al., 2021), it was anticipated that this variant would be significantly more resistant to neutralization by vaccinee sera. To experimentally validate this assumption, we generated pseudovirus with Omicron spike incorporating 34 distinct mutations (including three deletions and one insertion) relative to the Wuhan spike. Importantly, our spike did not contain R346K, which has been observed at relatively low frequency (<10%) within the Omicron lineage, but which is known to mediate escape from class 3 neutralizing monoclonal antibodies such as AZD1061/cilgavimab (Barnes et al., 2020; Dong et al., 2021; Greaney et al., 2021b).

As stated in our previous study, given the assumption of a polyclonal response to vaccination, we would have initially anticipated that small numbers of variations in spike protein would have only modest effects on recognition by the immune system (Garcia-Beltran et al., 2021a). However, our earlier findings suggested that the response to primary vaccination was relatively oligoclonal, hence providing a conceptual framework to explain how a relatively small number of mutations appears to mediate escape from humoral responses. Consistent with this framework, we found that the Omicron variant, harboring substantially more mutations than prior variants, efficiently escapes humoral immunity induced by primary vaccination. Strikingly, however, mRNA vaccine boosters appear to enable cross-neutralizing responses against Omicron, either by further affinity maturation of existing antibodies or targeting of new epitopes shared among variants.

Our study included distantly vaccinated individuals (>6 months) with evidence of prior infection. However, our approach could not distinguish between infection prior to vaccination or asymptomatic breakthrough infection. Irrespective of this distinction, these individuals exhibited higher levels of neutralizing activity against wild type, Delta, and Omicron pseudoviruses as compared to distantly vaccinated, infection-naive individuals. These results suggest that additional exposure to viral antigen through infection yields enhanced overall neutralization activity. It is important to note, however, that recent boosting in infection-naive individuals yielded substantially higher cross-neutralizing activity to Omicron as compared to prior infection.

Interestingly, vaccinated individuals who were boosted generated the broadest and most potent antibody responses of any cohort tested. This is surprising because the boost antigen was identical to the antigen used in the primary series, which would be expected to elicit memory responses that raise titers of the existing, strain-specific antibody repertoire. Samples from boosted individuals exhibited strong linear correlations between wild type and variant neutralization titers in contrast to non-boosted vaccinees, who exhibited a weak to absent correlation. These results suggest that neutralization titers achieved as a consequence of boosting resulted in humoral immunity with greater breadth than equivalent neutralization titers achieved with primary vaccination alone. Although the emergence of variants has catalyzed the development of variant-specific booster shots to increase variant neutralization, our results suggest that current wild type-based mRNA vaccines are sufficient to stimulate cross-reactive humoral responses greater than might have been anticipated. Whether this is the result of booster timing (in most cases >6 months after vaccination) or because the primary series was simply insufficient to fully stimulate B cell responses against each of the possible epitopes on the spike antigen is unclear.

We also found marked differences between approved vaccine regimens, with mRNA-1273 and BNT162b2 vaccines offering significantly greater humoral immune responses against all variants than Ad26.COV2.S, as has been previously described (Naranbhai et al., 2021a, 2021b). Recipients of mRNA boosting following Ad26.COV2.S exhibited markedly improved neutralizing titers, but did not achieve the breadth of response seen with mRNA primary vaccination followed by mRNA booster. Our results would suggest that these recipients of Ad26.COV2.S vaccines may benefit from additional mRNA vaccine doses with the potential to further raise titers and broaden their neutralizing activity.

Several reports have measured the decay of neutralizing antibody responses raised by mRNA COVID-19 vaccines with a half-life of 69-173 days (Doria-Rose et al., 2021; Levin et al., 2021). Our results from distantly vaccinated samples are in line with these reports given that neutralization activity against wild type pseudovirus was more than ten-fold lower in distantly vaccinated individuals. Given the activity generated against Omicron, it will be important to determine the longevity of cross-neutralizing antibody responses in boosted individuals as this could have a meaningful impact on vaccine efficacy as future variants of concern continue to emerge.

In support of the rapid spread of Omicron globally, our studies suggest that Omicron spike may exhibit increased infectivity. Interestingly, while the Beta variant previously exhibited escape from vaccine-induced humoral immunity, we found that infectivity of Beta was lower than wild type, perhaps explaining the relatively lower epidemic spread of Beta. In contrast, the now globally dominant Delta strain exhibited 2-fold increased pseudovirus infectivity relative to wild type, which may have contributed to its rapid spread. Additional studies examining the interaction between Omicron spike and ACE2 will be necessary to discern whether improved pseudovirus infectivity is also observed in replication competent virus.

Taken together, we demonstrate that Omicron drastically escapes vaccine-induced immunity after primary vaccination series with mRNA-1273 (Moderna), BNT162b2 (Pfizer-BioNTech) or Ad26.COV2.S (Johnson & Johnson/Janssen) and exhibits increased infectivity *in vitro*, raising the potential for increased transmissibility. Of note, despite escape from humoral immunity, Omicron breakthrough infections may result in attenuated disease severity in vaccinees due to cellular and innate immunity. However, neutralization remains the leading correlate of protection from infection, and this study demonstrates that receiving a third dose of an mRNA-based vaccine effectively yields a potent cross-neutralizing response against SARS-CoV-2 Omicron, likely through increasing breadth and cross-reactivity of neutralizing antibodies. These findings support the need for rapid and synchronized widespread deployment of mRNA boosters as a public health measure to curtail the emergence and spread of highly mutated SARS-CoV-2 variants.

### Limitations of the study

Although previous studies that use pseudovirus neutralization to model the sensitivity of replicating SARS-CoV-2 to neutralizing antibodies have shown excellent correlations (Crawford et al., 2020; Ju et al., 2020; Moore et al., 2004; Pinto et al., 2020; Riepler et al., 2020; Wang et al., 2020; Yang et al., 2020), it is possible that the mutations in Omicron spike protein may cause Omicron pseudovirus to behave differently than previously tested variants. However, recent reports have demonstrated similar loss of neutralizing activity by vaccinee sera against intact Omicron coronavirus (Cele et al., 2021b). In addition, while we confirmed that ACE2 expression is required for infection of 293T cells, natural target cells in the respiratory tract may express alternative receptors or attachment factors that facilitate infection and are not adequately modeled in our system. In addition, our cohort was cross-sectional and not longitudinal, which limits our ability to estimate changes in neutralization titers over time across single individuals. Furthermore, we did not assess other antibody-mediated functions such as complement deposition, antibody-dependent cellular cytotoxicity, or antibody-dependent cellular phagocytosis, which may contribute to protection even in the absence of neutralizing antibodies. We did not assess the role of vaccine-elicited cellular immune responses mediated by T cells and NK cells, which are likely to play a key role in disease prevention for vaccine recipients.

## Data Availability

All data produced in the present work are contained in the manuscript

## ACKNOWLEDGEMENTS, FUNDING SUPPORT

We wish to thank Michael Farzan, PhD for providing ACE2-expressing 293T cells. B.M.H. is supported by award Number T32GM007753 from the National Institute of General Medical Sciences. J.F. is supported by T32AI007245. D.J.G. and M.N.P. were supported by the VIC Innovation fund. A.G.S. was supported by NIH R01 AI146779 and a Massachusetts Consortium on Pathogenesis Readiness (MassCPR) grant. A.H. is supported by the *DZIF* (German Center for Infection Research, TTU 01.709). This work was supported by the Peter and Ann Lambertus Family Foundation. V.N. received support from a Medscape Young Investigators Lung Cancer award. A.B.B. was supported by the National Institutes for Drug Abuse (NIDA) Avenir New Innovator Award DP2DA040254, and the MGH Transformative Scholars Program. We thank Anand Dighe, MD and Andrea Nixon, BS, and the MGH Core Clinical laboratory for excellent assistance with clinical SARS-CoV-2 serology testing.

## AUTHOR CONTRIBUTIONS

W.F.G.B., K.S.D., E.C.L. and A.B.B. designed the experiments. W.F.G.B., K.S.D., E.C.L., V.N., A.D.N., A.R., and A.B.B. carried out experiments and analyzed data. W.F.G.B., V.N., C.B., O.O., C.C.C., D.G., M.C.P. and A.J.I led and performed cohort recruitment and sampling. J.F., B.M.H., and A.G.S. provided key reagents and useful discussions and insights. A.B.B., V.N., W.F.G.B., A.D.N., and A.H. contributed to statistical and sequence analyses. A.J.I. and V.N. provided key discussions and input into experimental design. W.F.G.B., V.N., A.H. and A.B.B. wrote the paper with contributions from all authors.

## MAIN FIGURE TITLES AND LEGENDS

**Table S1.** Clinical data from vaccinee cohorts, related to Figure 2. Clinical data is shown for each pre-defined cohort.

| Vaccinee cohort | recent vaccination<br>(<3 months) | distant vaccination<br>(6-12 months) | distant vaccination<br>+ infection | booster vaccination<br>(<3 months) |
| --- | --- | --- | --- | --- |
| **Total (n)** | **65** | **62** | **41** | **71** |
| **Demographics** |  |  |  |  |
| Age (y) (median, range) | 39 (24 – 72) | 41 (18 – 71) | 46 (22 – 78) | 34 (22 – 76) |
| Sex (% female) | 43/65 (66%) | 40/62 (65%) | 26/41 (63%) | 41/71 (58%) |
| Community cohort (%) | 20/65 (31%) | 40/62 (65%) | 33/41 (80%) | 11/71 (15%) |
| Healthcare worker cohort (%) | 45/65 (69%) | 22/62 (35%) | 8/41 (20%) | 60/71 (85%) |
| Days since last dose (median, range) | 17 (7 – 82) | 244 (181 – 321) | 229 (130 – 302) | 31 (7 – 118) |
| **mRNA-1273 (Moderna)** |  |  |  |  |
| **Total (n)** | **24** | **23** | **8** | **33** |
| Age (y) (median, range) | 54 (24 – 72) | 39 (23 – 65) | 36 (22 – 56) | 35 (23 – 74) |
| Sex (female, %) | 17/24 (71%) | 16/23 (70%) | 3/8 (38%) | 17/33 (52%) |
| Community cohort (%) | 0/24 (0%) | 9/23 (39%) | 4/8 (50%) | 3/33 (9%) |
| Healthcare worker cohort (%) | 24/24 (100%) | 14/23 (61%) | 4/8 (50%) | 30/33 (91%) |
| Days since last dose (median, range) | 18 (7 – 60) | 291 (181 – 315) | 243 (130 – 302) | 21 (7 – 82) |
| Cross-over vaccination regimen (%) | n.a. | n.a. | n.a. | 1/33 (3%) |
| **BNT162b2 (Pfizer)** |  |  |  |  |
| **Total (n)** | **21** | **33** | **27** | **30** |
| Age (y) (median, range) | 32 (25 – 65) | 43 (18 – 71) | 47 (22 – 78) | 33 (22 – 76) |
| Sex (female, %) | 13/21 (62%) | 22/33 (67%) | 18/27 (67%) | 19/30 (63%) |
| Community cohort (%) | 0/21 (0%) | 25/33 (76%) | 24/27 (89%) | 4/30 (13%) |
| Healthcare worker cohort (%) | 21/21 (100%) | 8/33 (24%) | 3/27 (11%) | 26/30 (87%) |
| Days since last dose (median, range) | 13 (7 – 32) | 224 (186 – 321) | 217 (180 – 300) | 49 (10 – 84) |
| Cross-over vaccination regimen (%) | n.a. | n.a. | n.a. | 6/30 (20%) |
| **Ad26.COV2.S (Janssen)** |  |  |  |  |
| **Total (n)** | **20** | **6** | **6** | **8** |
| Age (y) (median, range) | 46 (24 – 67) | 52 (34 – 71) | 44 (35 – 68) | 32 (23 – 37) |
| Sex (female, %) | 13/20 (65%) | 2/6 (33%) | 5/6 (83%) | 5/8 (63%) |
| Community cohort (%) | 20/20 (100%) | 6/6 (100%) | 5/6 (83%) | 4/8 (50%) |
| Healthcare worker cohort (%) | 0/20 (0%) | 0/6 (100%) | 1/6 (17%) | 4/8 (50%) |
| Days since last dose (median, range) | 50 (11 – 82) | 249 (184 – 269) | 247 (233 – 266) | 39 (20 – 118) |
| Cross-over vaccination regimen (%) | n.a. | n.a. | n.a. | 7/8 (88%) |

## STAR ★ METHODS

### RESOURCE AVAILABILITY

#### Lead Contact

Further information and requests for resources and reagents should be directed to and will be fulfilled by Alejandro Balazs and/or Wilfredo F. Garcia-Beltran.

#### Materials Availability

Plasmids generated in this study will be available through Addgene. Recombinant proteins and antibodies are available from their respective sources.

#### Data and Code Availability

This study did not generate sequence data or code. Data generated in the current study (including neutralization and antibody measreuments) have not been deposited in a public repository but are available from the corresponding author upon request.

### EXPERIMENTAL MODEL AND SUBJECT DETAILS

#### Human subjects

Use of human samples was approved by Partners Institutional Review Board (protocol 2020P002274). Serum samples from 239 COVID-19 vaccinees were collected. For each individual, basic demographic information including age and sex as well as any relevant COVID-19 history was obtained.

#### Cell lines

HEK293T cells (ATCC) were cultured in DMEM (Corning) containing 10% fetal bovine serum (VWR), and penicillin/streptomycin (Corning) at 37°C/5% CO2. 293T-ACE2 cells were a gift from Michael Farzan (Scripps Florida) and Nir Hacohen (Broad Institute) and were cultured under the same conditions. Confirmation of ACE2 expression in 293T-ACE2 cells was done via flow cytometry.

### METHOD DETAILS

#### Construction of variant spike expression plasmids

To create Delta and Omicron variant spike expression plasmids, we performed multiple PCR fragment amplifications utilizing oligonucleotides containing each desired mutation (Azenta) and utilized overlapping fragment assembly to generate the full complement of mutations for each strain. Importantly we generate these mutations in the context of our previously described codon-optimized SARS-CoV-2 spike expression plasmid harboring a deletion of the C-terminal 18 amino acids that we previously demonstrated to result in higher pseudovirus titers. Assembled fragments were inserted into NotI/XbaI digested pTwist-CMV-BetaGlobin-WPRE-Neo vector utilizing the In-Fusion HD Cloning Kit (Takara). All resulting plasmid DNA utilized in the study was verified by whole-plasmid deep sequencing (Illumina) to confirm the presence of only the intended mutations.

#### SARS-CoV-2 pseudovirus neutralization assay

To compare the neutralizing activity of vaccinee sera against coronaviruses, we produced lentiviral particles pseudotyped with different spike proteins as previously described (Garcia-Beltran et al., 2021b). Briefly, pseudoviruses were produced in 293T cells by PEI transfection of a lentiviral backbone encoding CMV-Luciferase-IRES-ZsGreen as well as lentiviral helper plasmids and each spike variant expression plasmid. Following collection and filtering, production was quantified by titering via flow cytometry on 293T-ACE2 cells. Neutralization assays and readout were performed on a Fluent Automated Workstation (Tecan) liquid handler using 384-well plates (Grenier). Three-fold serial dilutions ranging from 1:12 to 1:8,748 were performed for each serum sample before adding 50–250 infectious units of pseudovirus for 1 h. Subsequently, 293T-ACE2 cells containing polybrene were added to each well and incubated at 37°C/5% CO2 for 48 h. Following transduction, cells were lysed using a luciferin-containing buffer (Siebring-van Olst et al., 2013) and shaken for 5 min prior to quantitation of luciferase expression within 1 h of buffer addition using a Spectramax L luminometer (Molecular Devices). Percent neutralization was determined by subtracting background luminescence measured in cell control wells (cells only) from sample wells and dividing by virus control wells (virus and cells only). Data was analyzed using Graphpad Prism and NT50 values were calculated by taking the inverse of the 50% inhibitory concentration value for all samples with a neutralization value of 80% or higher at the highest concentration of serum. NT50 values were converted to WHO International Units using the Human SARS-CoV-2 Serology Standard (Lot #COVID-NS01097) obtained from NCI-Frederick National Laboratory for cancer research which was calibrated to the WHO SARS-CoV-2 Serology International Standard (20/136).

#### Titering

To determine the infectious units of pseudotyped lentiviral vectors, we plated 400,000 293T-ACE2 cells per well of a 12-well plate. 24 h later, three ten-fold serial dilutions of neat pseudovirus supernatant were made in 100 μL, which was then used to replace 100 μL of media on the plated cells. Cells were incubated for 48 h at 37°C/5% CO2 to allow for expression of ZsGreen reporter gene and harvested with Trypsin-EDTA (Corning). Cells were resuspended in PBS supplemented with 2% FBS (PBS+), and analyzed on a Stratedigm S1300Exi Flow Cytometer to determine the percentage of ZsGreen^+^ cells. Infectious units were calculated by determining the percentage of infected cells in wells exhibiting linear decreases in transduction and multiplying by the average number of cells per well determined at the initiation of the assay. At low MOI, each transduced ZsGreen^+^ cell was assumed to represent a single infectious unit.

#### Quantitation of pseudovirus by RT-qPCR

To determine the genome copy concentration of pseudotyped lentiviral vectors, lentiviral RNA was extracted from pseudovirus supernatant using the QIAamp viral RNA mini kit (Qiagen). Each sample was serially diluted, and each dilution was treated with 1.2 U of Turbo DNase (Invitrogen) at 37°C for 30 min followed by heat inactivation at 75°C for 15 min. 10 μL of the treated RNA was used in a 20 μL qRT-PCR reaction with the qScript XLT one-step RT-qPCR Tough Mix, low ROX mix (Quanta Biosciences), a TaqMan probe containing locked nucleic acids (/56-FAM/AGC+C/i5NitInd/GG+GA/ZEN/GCTCTCTGGC/3IABkFQ/) (IDT), and primers designed targeting the LTR gene of NL4-3 HIV genome, from which the lentiviral vector was derived (5’-GGTCTCTCTIGITAGACCAG and 3’-TTTATTGAGGCTTAAGCAGTGGG). Each dilution was run in duplicate on a QuantStudio 12K Flex (Applied Biosystems). The following cycling conditions were used: 50°C for 10 min, 95°C for 3 min followed by 50 cycles of 95°C for 15 s and 60°C for 1 min. Virus titer was determined by comparison with a standard curve generated using a plasmid standard generated from serial dilution of CMV-Luciferase-IRES-ZsGreen lentiviral backbone. DNase and No DNase controls were also included at 2.5 × 10^8^ GC/mL of the same plasmid. The range of the assay was from 2.5 × 10^7^ GC/mL to 1.5 × 10^3^ GC/mL. Upon analysis, the average of the three most concentrated dilutions within range of the standard were used to calculate genome copies/mL.

#### Anti-SARS-CoV-2 spike and nucleocapsid antibody assays

The EUA-approved electrochemiluminescence-based Roche Elecsys® anti-SARS-CoV-2 spike antigen (semi-quantitative) and nucleocapsid antigen (qualitative) immunoassays were used to detect total antibodies (IgG, IgM, and/or IgA antibodies) to SARS-CoV-2 spike and nucleocapsid in vaccinee sera. The upper limit of detection for the spike semi-quantitative assay was 25,000 U/mL. The assay was run on a Roche Cobas 8000 e801 Immunoassay Analyzer in the Massachusetts General Hospital Core Laboratory.

### QUANTIFICATION AND STATISTICAL ANALYSIS

Data and statistical analyses were performed using GraphPad Prism 9.2.0, JMP Pro 16.1.0 (SAS Institute), and R v4.0.2. Flow cytometry data was analyzed using FlowJo 10.7.1. Student *t* tests were performed in the indicated figures; all *p* values are unadjusted. Statistical significance was defined as *p* < 0.05. Error bars throughout all figures represent one standard deviation or standard error of the mean where indicated.

